# The lived experience of rejection sensitivity in ADHD - a qualitative exploration

**DOI:** 10.1101/2024.11.16.24317418

**Authors:** Annabel Rowney-Smith, Beth Sutton, Lisa Quadt, Jessica A Eccles

## Abstract

Attention Deficit Hyperactivity Disorder (ADHD) is a neurodevelopmental condition, commonly associated with differences in attention, and/or hyperactivity and impulsiveness. A lesser known, but highly impactful characteristic of ADHD is emotional dysregulation, which causes difficulties in emotional expression and identification. An aspect of emotional dysregulation which remains relatively unexplored in ADHD, is rejection sensitivity. In people who experience rejection sensitivity, perceived rejection and/or criticism can evoke extreme dysphoria. To investigate the lived experience of rejection sensitivity in ADHD individuals, five undergraduates participated in focus-group interviews. The subsequent thematic analysis revealed three key themes which encapsulated the experience; *withdrawal*, *masking*, and *bodily sensations*. The participants explained how rejection sensitivity elicited unpleasant bodily sensations, anxiety, and misery, and how, in turn, they used masking to camouflage these feelings. They then discussed how the use of masking had caused them to become dissociated from themselves and to withdraw from others, which often resulted in loneliness. It was apparent that rejection sensitivity significantly impacts mental wellbeing, eliciting feelings such as anxiousness, despair, and embarrassment. In turn, social function is significantly impaired, alongside career opportunities, and daily life. Our findings indicate a need for deeper understanding of ADHD traits and emotional dysregulation, which may in turn lessen the presence of rejection sensitivity in ADHD individuals.

## Introduction

ADHD is a neurodevelopmental condition, characterised by differences in attention and emotion regulation, possible hyperactivity, and impulsiveness (1). The neurodiversity movement (2) advocates that neurodevelopmental conditions are often inseparable from one’s identity (3). Thus, to align our research with the neurodiversity movement’s values, the language in this paper is henceforth adapted to avoid the perpetuation of ableist stances. For example, medicalised language will be avoided, e.g., ‘traits’ instead of ‘symptoms’, and identity-first language will be used instead of person-first language (e.g., ‘ADHD person’, or ‘ADHDer’, instead of ‘person with ADHD’).

Emotional dysregulation is overrepresented in ADHD and may be prevalent in up to 70% of ADHD adults (4). emotional dysregulation is multi-dimensional, but often presents as increased and/or reduced emotional reactivity, lack of emotional awareness, and intense emotional expression (5). Consequently, ADHDers may experience an array of emotional dysregulation-related symptoms, including intense mood swings, and reactivity to seemingly benign situations. Thus, emotional dysregulation is thought to significantly contribute to psycho-social problems more often faced by ADHD people, such as unemployment and interpersonal issues (6) and has been found to be more negatively influential on quality of life than inattentive and hyperactivity traits (7). Despite its common association with ADHD, and its significant effects, emotional dysregulation remains an often-neglected trait, which is evidenced in its exclusion from diagnostic criteria (8). In recent years, anecdotal reports have highlighted a crucial dimension of emotional dysregulation experienced by ADHD people; rejection sensitivity (9).

Rejection sensitivity is defined as dysphoria induced by perceived or real rejection, and/or criticism (9). People who experience rejection sensitivity readily detect rejection, and therefore perceive rejection/criticism more frequently than others (10). The ensuing dysphoric episode often involves intense feelings of extreme misery, anxiety, and pain (11). The source of rejection sensitivity is disputed, however; evidence suggests that childhood interpersonal trauma, such as caregiver rejection, may contribute to causality (12). Although rejection sensitivity is defined as a personality disposition (13), it frequently features in mental health disorders typically characterised by emotional dysregulation, such as depression, Body Dysmorphic Disorder, and Borderline Personality Disorder (14).

To date, few studies have investigated the potential association between ADHD and rejection sensitivity. Moreover, these studies, for the most part, have attempted to establish the *extent* of the rejection sensitivity -ADHD association, rather than explore the *nature* of the association. However, although they did not directly measure rejection sensitivity, one study (15) explored the experience of received criticism in an ADHD population, via qualitative techniques (open-ended written responses).

Since emotional dysregulation presents as a significant influence on quality of life for ADHDers, and rejection sensitivity may be a contributing factor in this, there is an obvious need for more research exploring the experience of rejection sensitivity in ADHD cohorts. Motivated by this gap in existing research, our study aimed to provide rich accounts of rejection sensitivity in an ADHD population, via qualitative methods; focus-group interviews and subsequent thematic analysis. We hope that this research will improve the understanding of neurodivergent experiences and shed light on the co-occurrence between rejection sensitivity and ADHD.

## Methods

### Measures

To explore the lived experience of rejection sensitivity, a qualitative study design was utilised; focus groups interviews. This approach was selected with the aim of capturing a rich, and illustrative collection of perspectives, as participants can explore and discuss their experiences alongside like-minded peers, and with limited restriction. At the start of the focus groups, informed verbal consent was gained from the participants, and time was allocated for introductions. The interviews were lightly structured by a topic guide which comprised one broad, open-ended question with associated sub-questions. The broad, over-arching question was aimed to frame the ensuing discourse; *“If you do, how do you identify with Rejection Sensitive Dysphoria?”*; whereas the associated sub-questions were intended to encourage participants’ engagement with the topic, and included questions such as, *“Has rejection sensitivity D interfered with your daily life?”*, and *“Do you experience any particular feelings when you are experiencing rejection sensitivity D?”.* To increase accessibility, the focus groups were conducted via online video-call, through Zoom.

### Participants

Five undergraduate students were recruited from the University of Sussex, via flyers distributed across the university campus. To participate in the study, participants had to satisfy inclusion/exclusion criteria; all participants were undergraduates with formal ADHD diagnoses and were over 18 years old. The participants were also incentivised to take part, via a £16 GiftPay voucher. Two focus group interviews took place, whereby one group contained two participants, and one group contained three. Participants were sent a copy of the consent form and an information sheet prior to providing verbal consent, to allow adequate understanding of the study expectations and the research process.

### Analysis

The focus group content was audio-recorded via a Dictaphone and transcribed verbatim into written text. Following this, the transcribed data was analysed and interpreted, informed by Braun and Clarke’s (16) six-step thematic analysis framework (16). To promote familiarity with the data, the focus-group transcriptions were firstly read multiple times. Next, seemingly important data were identified, and then organised into themes according to patterns of similarity. The potential themes were then discussed and reviewed amongst the research team, whereby three defined themes emerged; *masking*, *withdrawal*, and *bodily sensations*. Two themes (*problems with self-esteem* and *cognitions*) were rejected because they overlapped with the other themes, and were determined as subthemes of our three defined themes.

Our study method was devised by University of Sussex researchers and undergraduate students alike, with neurodivergent perspectives.

Ethical approval for this study was obtained by the Research Governance and Ethics Committee at Brighton and Sussex Medical School (application no. ER/AR649/1). Data was collected between 25^th^ July 2022 and 2^nd^ September 2022.

## Results

Participants spoke in depth about their feelings, thought processes, and physical experiences before, during, and after rejection/criticism. In commonality, they described rejection sensitivity as a painful and overwhelming experience, which often lasts a significant amount of time (between hours and weeks) and can resurface many years later. Three primary themes emerged from the data; *masking*, *withdrawal*, and *bodily sensations* (Fig. 1).

**Figure 1.**
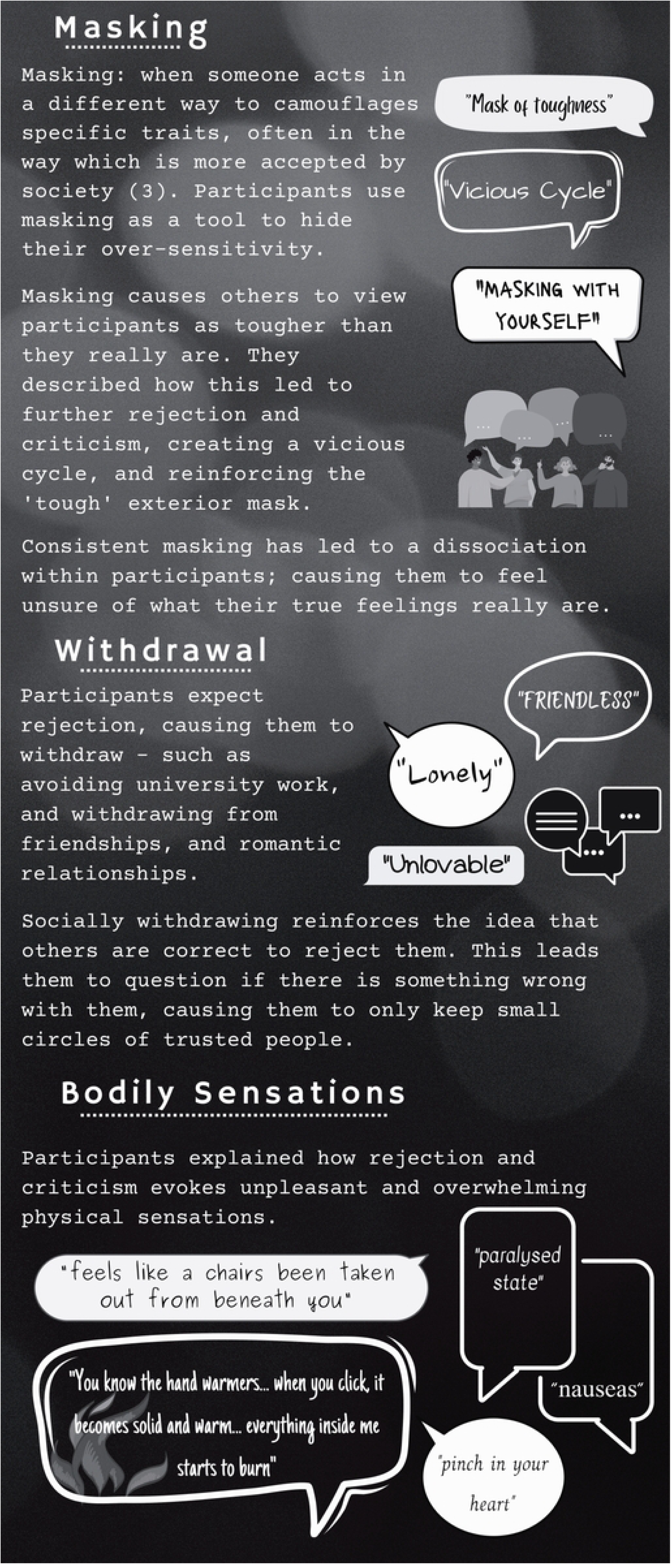
Graphic depicting the key themes in the analysis of rejection sensitivity in ADHD

### Withdrawal

One of the most common themes identified was ‘withdrawal’. The participants explained that rejection sensitivity often caused them to withdraw from friendships, family and romantic relationships, university life, and job opportunities. All participants agreed that the *expectation* of rejection elicited more dysphoria than the rejection *itself*; “there’s a general idea that I will be rejected”. Thus, participants reported to pre-emptively withdraw to avoid potential rejection; “I just get myself out of the situation where the rejection might come, just to avoid it”.

Participants discussed how withdrawal negatively impacts their relationships with friends and family. It was disclosed that they “rarely reach out” to people, and that when they *do* contact people, they consistently assess for possible rejection/criticism. For instance, they may assume that an unanswered text message is a rejection; “I just ended up being awake until four in the morning… like constantly checking my phone, being like, why was this person not responding?”. Therefore, participants explained that it is easier to avoid contacting people, as this pre-emptively lessen rejection sensitivity anxiety. In turn, their quantity and quality of relationships is negatively impacted. One participant expressed that there were “limited people in my life that I’m close with”. Similarly, another participant shared that they keep their “friendship circles low and full of neurodivergent people” as they had suffered “so much rejection from allistic people”. Consequently, participants disclosed how they often felt “lonely”, “friendless”, and “unlovable”.

Withdrawal also affects participants’ university work; most participants agreed that the fear of rejection/criticism had caused them to delay, or not submit assignments. A few participants admitted that in order to protect themselves from criticism, they had purposefully submitted university work which was below their usual standard, or had submitted it late (which typically incurs a 10% grade deduction). They explained that this allowed them an explanation for potential rejection/criticism in the form of a disappointing grade. Focus group members also discussed withdrawal from the workplace. One participant had avoided applying for career opportunities “to avoid rejection”. Interestingly, participants explained how they often experienced rejection sensitivity during the application process, as opposed to when they *received* the rejection. This conforms with the earlier assertion, that the expectation of rejection “is more damaging than the actual rejection”. Participants discussed that they do not feel they deserve good university grades or successful job applications, with one participant employing the term; “imposter syndrome”.

### Masking

Masking is often utilised by neurodivergent people to suppress and camouflage neurodivergent traits (17). This is achieved via the employment of multiple tools (both consciously and sub-consciously), and subsequent alterations in behaviour (17). Most participants, either explicitly or implicitly, used masking to camouflage their thoughts and feelings about a perceived rejection/critique. The participants discussed their individual difficulties with distinguishing between “real” and their perceived rejection. A participant provided an example of this; they explained that they could not differentiate between jokes and critiques. Thus, participants explained that they often felt the need to ask friends/family for clarification and/or reassurance. However, this often led to embarrassment, anxiety, invalidation, and labels such as “oversensitive”. To avoid this, participants mask to convey robustness, and a nonchalant attitude to rejection; “a mask of toughness”. This enables them to hide their rejection sensitivity from others and helps them feel less like an outsider, and while it helps them temporarily feel like an outsider, participants agreed that masking has negative consequences.

Participants revealed that consistent masking causes disconnection from their emotions and their authentic selves, as they struggle to distinguish between real and faux feelings; “I think at some point it’s not just masking towards others, but you are also masking with yourself… you can’t identify if you’re bothered or not, because you lose connection”. Participants also described how masking caused them to feel disconnected from social interaction; “observing this interaction that’s happening to you”. One participant disclosed how they had benefitted from therapy, because it enabled them to identify and reconnect with their emotions. The participants also explained that masking perpetuates their social withdrawal. Two participants described this as a “vicious circle”, whereby others take advantage of their nonchalant mask, and often incorrectly assume that rejection and criticism does not affect them. Consequently, rejections and critiques increase, and they subsequently withdraw further from social situations, and further reinforce the mask; a vicious circle.

### Bodily Sensations

Participants described how perceived rejection evokes overwhelming and unpleasant bodily sensations. Interestingly, sensations were varied and unique to the individual. For example, one participant felt sensations in their heart and throat; “it’s like a pinch in your heart, like a very swift, brief, like, like your heart or like your throat closing up… feels like your heart kind of drops maybe a bit”. Another participant explained how they felt as if “a chair (had) been taken out from beneath you”. One participant used a powerful metaphor to explain the flooding of heat throughout their body; “you know the hand warmers, when you click, it becomes solid and warm… getting warmer inside me…. everything inside me starts to burn”. In contrast, a participant discussed how they felt their body enter a “paralysed state”. Stomach sensations were the only sensation common amongst participants; “sick to your stomach”, and nausea “in my gut”.

## Discussion

Our findings indicate that rejection sensitivity is a multidimensional experience; encompassing emotional pain, negative cognitions, and unpleasant physical sensations. Our participants explained how the effects of rejection sensitivity, like anxiety, depression, and unpleasant bodily sensations, had led them to mask their true feelings, and withdraw from relationships and work/university commitments. It was apparent that rejection sensitivity is detrimental to mental health, and impairs both daily function and long-term wellbeing.

The findings also indicate that rejection sensitivity could be a contributing factor in the psychosocial problems often faced by ADHDers. For example, the participants explained how they are less likely to apply for jobs or submit university assignments because they were expecting rejection and were protecting themselves from the ensuing misery of rejection sensitivity. This concern could potentially limit their job prospects, and any future career aspirations. Furthermore, subsequent withdrawal from relationships may restrict opportunities to find a romantic partner and friends, and in extreme cases, could lead to familial estrangement. Again, this could contribute to psychosocial problems, as ADHDers may find themselves without a support network, and may find it hard to start families of their own. Thus, the effects of rejection sensitivity are far-reaching, and could prove long-term. Previous research has primarily focussed on how ADHD traits affect psychosocial functioning, yet our findings suggest that rejection sensitivity may be just as important in the development of psychosocial issues.

The participants spoke at length about how their withdrawal from friends and family had left them lonely and isolated. Research has found that ADHDers are more likely to experience loneliness than those without ADHD (18), and rejection sensitivity - masking may contribute to this. Moreover, research also indicates that masking threatens the development and maintenance of deep relationships. Bradley et al. found that masking made their autistic participants feel disingenuous because their relationships were based on an untrue representation of themselves (19). Their participants felt that their relationships with others were based on deception, and that very few people truly knew who they truly were. This is echoed in our study, when our participants expressed how they use masking to present themselves as tougher than they are, which, in turn, made others feel as if they could continue to exploit them, not knowing the devastating consequences for the participants’ mental health. This may further contribute to the participants’ withdrawal from relationships with others, as they may find their relationships unfulfilling, shallow, and based on deception. A study conducted by Houghton et al. (20) corroborates this, as they found that ADHDers often report their friendships as low quality (20). They also found that loneliness mediates the often-co-occurrent depressive disorders in ADHD (20). rejection sensitivity could form part of the driving force behind the high prevalence of loneliness and depression in ADHD people, as it may cause ADHDers to have superficial friendships, and withdraw from developing and maintaining relationships.

Our participants disclosed how their extensive use of masking had caused them to feel dissociated from themselves; one participant explained how they needed therapy to re-identify their true self. Dissociation is often allied with mental health disorders like bipolar disorder (21) and borderline personality disorder (BPD) (22), which are both common ADHD-co-occurrent conditions (23, 24). It may be that rejection sensitivity -masking contributes to the development of bipolar disorder and/or BPD in ADHDers through its dissociative consequences. For instance, our participants discussed how they felt that masking had led them astray from their true identities, leading them on to question who they really are. This unstable self-image and de-personalisation is also frequently seen in people with bipolar disorder and BPD. Furthermore, since BPD sufferers are more likely to experience unstable relationships (25), rejection sensitivity -related withdrawal from relationships may *also* contribute to BPD symptoms. Unfortunately, due to symptoms overlap (primarily emotional dysregulation-related), ADHD is often misdiagnosed as BPD, especially in females (26). It may be that either rejection sensitivity -masking and withdrawal contributes to the *development* of BPD in ADHDers, or they may also be distinct features of ADHD, which would contribute to the *misdiagnosis* of BPD.

To our knowledge, the unpleasant bodily sensations felt by the participants was a new finding in the context of rejection sensitivity. We found that these sensations varied greatly amongst the participants; each participant experienced different physical sensations in different bodily areas. For example, one participant primarily felt nausea in their stomach and gut, whereas another felt that their heart and throat were tightening. Additionally, we found that rejection sensitivity affects the perception of bodily temperature, when one participant explained that rejection sensitivity evoked the feeling of progressively intense heat throughout their body. Conversely, rejection sensitivity also appeared to *inhibit* the feeling of physical sensation, when one participant explained that rejection sensitivity causes them to feel paralysed. These types of bodily sensations are often found in anxiety disorders, and may reflect an anxiety attack. Anxiety attacks are caused by activation of the sympathetic nervous system, which effectively causes the body to enter a fight-or-flight physiological state (27 p. 189–190). This affects multiple bodily systems, often resulting in various symptoms, such as nausea, feelings of warmth or cold, chest tightness, and abdominal pain (27 p. 189–190). It may be that rejection sensitivity elicits a stress response in ADHDers, which may contribute to the often co-occurrent diagnosis of anxiety disorders.

Perhaps the most insightful and important notion to arise from our participants’ honesty, is the idea that rejection sensitivity could be prevented or alleviated if they received more understanding and empathy from others. The participants expressed that they consistently felt they needed reassurance from others to be able to help alleviate rejection sensitivity symptoms. They explained that if they did not receive the reassurance they needed, their rejection sensitivity -related anxiety would “spiral”, because they would repeatedly analyse the rejection sensitivity -provoking situation; trying to work out if they had been rejected or not. However, they often felt embarrassed to ask for reassurance, because they did not want to present as annoying or as overly sensitive. This was mainly due to previous negative experiences, whereby asking for reassurance was followed by judgement from others. Thus, gaining reassurance was a *double-edged sword*; seeking reassurance induced feelings of embarrassment and judgement, and alternatively, not seeking reassurance induced anxiety. This may also contribute to withdrawal from relationships, as participants may wish to avoid the consequences of either decision. This finding indicates that more understanding and empathy from others may mediate the severity of rejection sensitivity, whereby ADHDers would not need to mask as much, and would potentially be able to socialise more without such negative consequences for their mental health.

Although our findings provide a rich insight into the experience of rejection sensitivity, they do not elucidate the aetiology of rejection sensitivity. However, it may be that rejection sensitivity develops as a result of sensitisation to sustained criticism. Due to lack of knowledge and understanding of trait aetiology, ADHD is classically stigmatised (28). ADHD traits may contravene social norms and can appear to others as if they are controllable. For example, neurotypical people are typically able to control their impulses, such as not interrupting others. This, paired with misconceptions regarding aetiology, lead to discreditation and prejudice (28). Thus, ADHD traits are often met with criticism. Since ADHD is present from childhood, ADHD people are more frequently exposed to criticism and rejection from a young age than neurotypical peers (29). It could be hypothesised that rejection sensitivity results from sensitisation; whereby the response to upsetting, frequent, and early experiences of criticism and rejection, become larger and more intense over time. This fits with the data collected from our focus-groups; the participants often felt that the anticipation of rejection was more painful than the rejection itself, indicating that past experience of rejection frames future response. This correlates with the increasing emotional dysregulation seen from childhood ADHD into adulthood ADHD (4). This sensitisation theory is corroborated by Babinski et al. (30), who investigated social feedback circuitry in adolescents with both ADHD and rejection sensitivity. The authors found that rejection sensitivity symptoms correlated with sensitisation of attentional neural circuitry for social rejection, and with desensitisation for positive social feedback (30). Therefore, ADHD people who experience rejection sensitivity, may be *oversensitive* to rejection cues, and *desensitised* to social approval cues, due to alterations in neural connections.

Consequently, rejection is perceived more frequently, and is unassuaged by *missed* social approval cues. Since ADHD is characterised by differences in attention, it may be that ADHD neuronal networks are more susceptible to this attentional imbalance between rejection and approval cues. Therefore, a combination of frequent and early rejection/criticism, paired with attentional wiring differences, may make ADHDers more vulnerable to rejection sensitivity.

Our findings aligned with previous literature, which found significant associations between rejection sensitivity and ADHD, with increased aggression and social anxiety (31, 32), higher levels of victim justice sensitivity and lower levels of perpetrator sensitivity (33), also withdrawal from social situations and relationships (32, 34). Although a previous study investigated the experience of criticism in people with ADHD traits (29), our study was, to the best of our knowledge, the first to directly explore the experience of *rejection sensitivity* in ADHD individuals.

Although our method was successful for permitting rich and illustrative descriptions of the experience of rejection sensitivity, the sample size was small, and we did not screen for other neurodivergent conditions. During the interviews, two participants disclosed co-occurring Autism Spectrum Condition diagnoses, whilst another disclosed a Dyslexia diagnosis. Since ADHD is frequently accompanied by other conditions, participants may have had other diagnoses we were unaware of. Future research may be disposed to use a larger sample, and to screen for other conditions, in order to improve generalisability and validity. This may also elucidate the possibility that ADHD mediates associations between rejection sensitivity and other conditions which are also co-occurrent with ADHD, such as BPD. We hope that our findings improve understanding of neurodivergent experiences and highlight the need for further research into potential underlying mechanisms.

## Acknowledgments

We thank all of our participants for their time and openness.

## Ethical approval statement

All elements of this study were approved by the Brighton & Sussex Medical School Research Governance & Ethics Committee (approval no. ER/AR649/1) on July 10^th^, 2022. The participants provided informed verbal consent before taking part.

## Data availability statement

All relevant data are within the manuscript. Data cannot be shared publicly because we have no permission from the respondents of our study to share the de-identified dataset with the general public (Ethics committee: Research Governance and Ethics Committee at Brighton and Sussex Medical School (application no. ER/AR649/1). Data requests can be directed to the corresponding author.

